# Systematic Review Protocol of aetiology of mechanical bowel obstruction in Low-and-middle income countries: Has anything changed in the last two decades?

**DOI:** 10.1101/2023.12.05.23299503

**Authors:** Yakubu Kevin Kwarshak, Mohammed Nakodi Yisa, Oghenegare Asheaba Kigbu, Daniel Akut John, Nankam David Jimwan, Karen Chineme Ubabuike, Peter Mkurtar Yawe

## Abstract

**Background:** Despite various causes of mechanical obstruction, there appears to be a great deal of variation depending on geographical location and age. Geographically, postoperative adhesions and hernia have been documented as the most common aetiology of mechanical bowel obstruction in high-income and low-and-middle-income countries, respectively. Whether there has been a change in this trend in low- and middle-income countries is a matter of speculation in the surgical community. Therefore, to fill this knowledge gap, this study aims to systematically review the existing literature on the aetiology of mechanical bowel obstruction with a focus on understanding the most common cause of mechanical bowel obstruction in low- and middle-income countries in both paediatric and adult populations to guide surgical practice.

**Methodology and Analysis:** This protocol was designed and written according to the guidelines of the Preferred Reporting Items for Systematic Review and Meta-analysis Protocol 2015 (PRISMA-P 2015) statement. However, the results of the systematic review will be reported in accordance with the Preferred Reporting Items for Systematic Review and Meta-analysis (PRISMA) statement. We will consider studies published in English and French between 2002 and 2022 that reported on the aetiology of mechanical bowel obstruction in any age group in low- and middle-income countries. We will conduct a literature search using Ovid MEDLINE, Ovid Embase, CINAHL on EBSCO and Web of Science databases employing relevant subject headings, keywords and synonyms, which will be combined using Boolean operators to refine the search results. A hand search of references of retrieved literature will be conducted. The retrieved articles will be imported into Zotero for de- duplication. The resulting set of titles and abstracts will be uploaded to Rayyan (an AI-assisted online systematic review tool), where they will be double-checked to identify articles eligible for inclusion. Two independent reviewers will screen articles to be included and disagreement will be resolved by discussion or by a third reviewer as a tie-breaker. Also, data extraction will be done by one reviewer and confirmed by another. Critical appraisal to assess the quality of the included studies will be carried out by two independent reviewers using the Joanna Briggs Institute (JBI) tools. We anticipate that the eligible studies will be quite heterogeneous in terms of their design, outcomes of interest, populations and comorbidities. Therefore, results may be synthesised descriptively without meta-analysis using charts, graphs and tables. Where possible, we will conduct a sub-analysis using conceptual frameworks based on age, WHO regions and continents.

**Ethics and Dissemination:** No ethical approval will be sought because the required data is already in the public domain. Findings will be published in peer-reviewed journals.

## INTRODUCTION

Acute bowel obstruction is one of the most common surgical emergencies encountered by surgeons (1), particularly by surgical trainees in the Emergency Department (ED). It accounts for 3% of all emergency admissions and 15% of cases of acute abdominal pain (1). More than two-thirds of bowel obstructions are mechanical small bowel obstructions (2). Mechanical bowel obstruction is an obstruction to the forward movement of bowel contents caused by a variety of causes and is characterised by a combination of the sudden onset of vomiting, abdominal pain, distension and constipation, with a high likelihood of strangulation and/or gangrene if not diagnosed and treated promptly (1–5) Therefore, this disease tends to be associated with high morbidity and mortality, especially in low-and-middle-income countries (LMICs) where the majority of patients are seen in secondary care settings. Several papers have reported a mortality rate of 5-13.5% associated with this mechanical obstruction, which is higher in LMICs (3,5,6). Preventing this mortality requires an up-to-date understanding of the common causes of this surgical pathology so that surgeons and trainees can make a correct and timely diagnosis and take appropriate therapeutic measures.

Mechanical bowel obstruction has many causes. Post-operative adhesions have been reported as one of the most common aetiologies. They are reported to occur in approximately 93% of patients who have undergone abdominopelvic surgery (1). The most commonly observed sources of peritoneal adhesions are appendectomy, gynaecological surgery and colorectal surgery (1,3). However, only 5% of these are symptomatic(1). Another important and common cause of bowel obstruction is hernia, such as inguinal, umbilical and incisional subtypes (4). Other aetiologies include volvulus, intussusception, intestinal malignancy, vermiform impaction, Meckel’s diverticulum and intestinal tuberculosis (3,4,7).

Despite these various causes of mechanical obstruction, there appears to be a great deal of variation depending on geographical location and age (8). In terms of age, hernia and adhesion as well as anorectal malformation and intussusception have been reported to be the most common causes in adults and children, respectively (1,3,8,9). Geographically, postoperative adhesions have been documented as the most common aetiology of mechanical bowel obstruction in high-income countries (HICs). This may be due to the high volume of abdominal surgery (8,9). In contrast, many studies have reported hernias as the most common aetiology in LMICs (1,4,8,10).

Whether there has been a change in this trend in LMICs is a matter of speculation in the surgical community. Although there are international empirical studies that have evaluated the changing trend in the causes of mechanical bowel obstruction in different countries of the LMICs, the results of these studies are contradictory (2–5,7,10,11). While some showed that hernia remained the most common cause in LMICs (1,3,4,8), others showed changing patterns towards postoperative adhesions (2,5,11). As a result, this review intends to draw together the overall body of evidence to guide safe surgical practice. In addition, most of these empirical studies are retrospective and characterised by a low level of evidence. They need to be reviewed to generate higher-level evidence. To the best of our knowledge, no systematic review has been conducted in all LMICs to assess the aetiological pattern of mechanical bowel obstruction, leaving a lack of up-to-date evidence to guide surgical trainees and surgeons in the prompt diagnosis and management of such an important surgical emergency. Therefore, to fill this knowledge gap, this study aims to systematically review the existing literature on the aetiology of mechanical bowel obstruction with a focus on understanding the most common cause of mechanical bowel obstruction in LMICs in both paediatric and adult populations to guide surgical practice. Given the high morbidity and mortality associated with this condition, understanding the common aetiological pattern of mechanical bowel obstruction is critical to improving overall outcomes, as this would help surgical trainees in the emergency department to maintain a high index of suspicion and avoid delays in diagnosis, referral or appropriate prompt intervention. This is particularly important given the fragile health systems and low numbers of surgeons in LMICs.

## METHODOLOGY

This protocol was designed and written according to the guidelines of the Preferred Reporting Items for Systematic Review and Meta-analysis Protocol 2015 (PRISMA-P 2015) statement(12). This framework prescribes best practices for the development and reporting of systematic review protocols. The review has been registered in PROSPERO, an international digital repository for the pre-registration of systematic reviews, identified by CRD42023468901. There are several benefits to registering the research protocol, including avoiding redundant research efforts, reducing bias, and increasing the overall transparency of the review process. The results of the systematic review will be presented in accordance with the Preferred Reporting Items for Systematic Review and Meta-analysis (PRISMA) statement(13). Should any deviations from the established protocol occur during the conduct of the review, these will be duly accounted for in the final publication.

### Eligibility

We will consider studies published between 2002 and 2022 in low- and middle-income countries (LMICs) that are either case series, case-control or cohort studies reporting on the aetiology of mechanical small bowel obstruction in any age group. Studies must be published in either English or French. We will exclude animal studies, studies with different designs, those published in high-income countries, those outside the specified date range, or those published in languages other than English or French. Should our manuscript be accepted for publication, an updated literature search will be conducted by employing the peer review study methodology. This will prevent missing on important new evidence.

### Data Sources

We will conduct a literature search using Ovid MEDLINE, Ovid Embase, CINAHL on EBSCO and Web of Science databases. Our search strategy will include the use of relevant subject headings, keywords and synonyms, which will be combined using Boolean operators to refine the search results. To focus our search on low- and middle-income countries (LMICs), we will use the ScHARR geographic filter (14) to exclude studies from non-LMIC countries. In addition, we will manually search the reference lists of the included studies to find additional relevant articles.

### Search Strategy

A search strategy was developed using the key terms ‘small bowel obstruction’, ‘lower-middle-income countries’ and ‘aetiology’. The MEDLINE search strategy, adapted for other databases, is shown in Figure 1.

**Figure 1:**
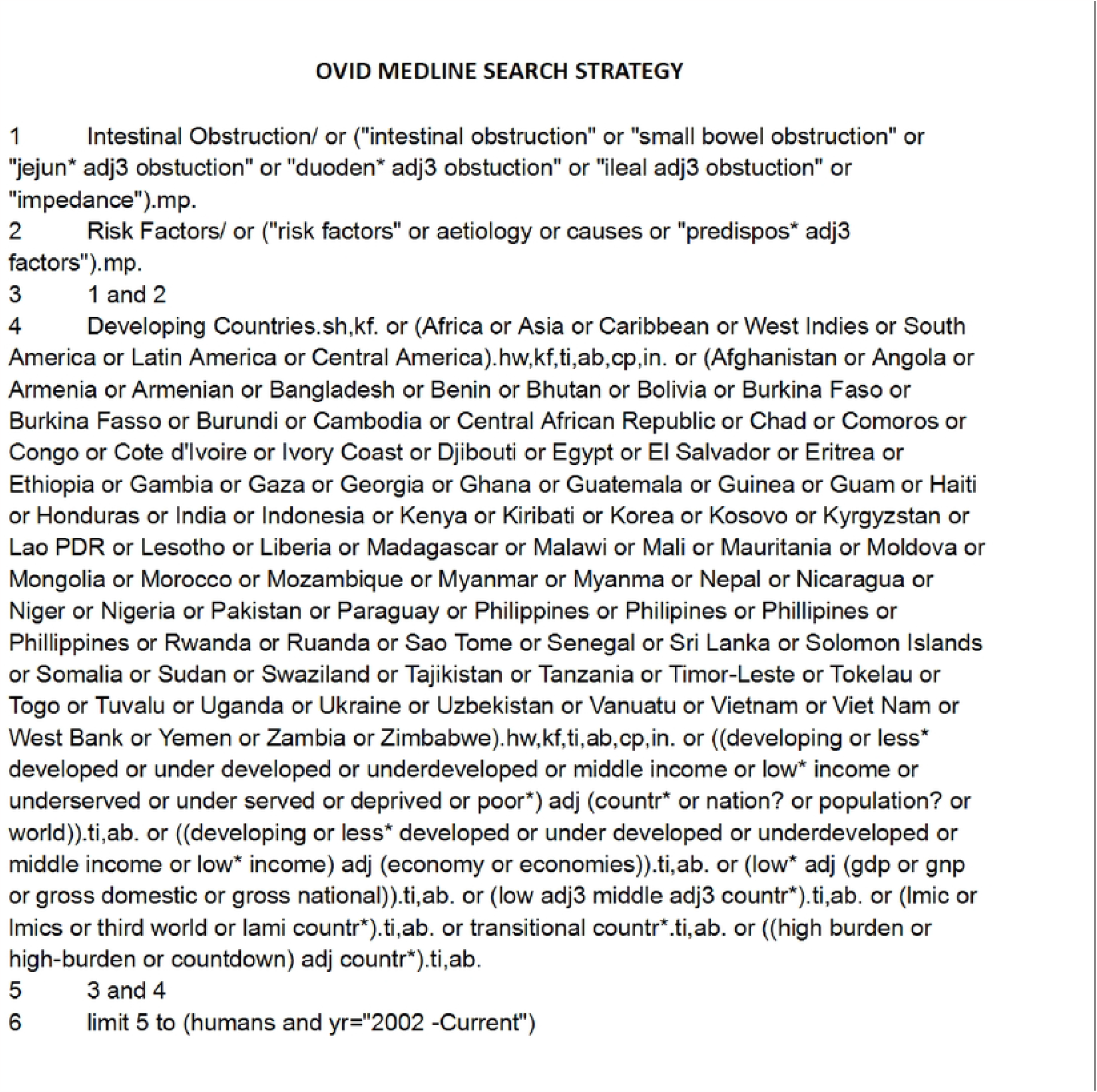
MEDLINE search strategy

### Study selection and data extraction

The retrieved references will be imported into Zotero for de-duplication. The resulting set of titles and abstracts will be uploaded to Rayyan (15) (an AI-assisted online systematic review tool), where they will be double-checked to identify articles eligible for inclusion. These will then be exported to Zotero for reference management and full-text screening. Studies that do not meet the eligibility criteria at this stage will be excluded, with reasons documented. This process is carried out by two independent authors. Disagreement will be resolved by discussion or by a third reviewer as a tie-breaker.

A data extraction form adapted from the JBI will be used for data extraction. The variables to be extracted from the included studies are described in Table 1. The data extraction form will be pre-tested independently by two reviewers using randomly selected articles. This will help to identify gaps that can then be used to refine the form. Data extraction will be carried out by two reviewers; reviewer A will extract the data, while reviewer B will check the accuracy of the extracted data

**Table 1:** List of items to be extracted from included studies.

| S/N | Item |
| --- | --- |
| 1. | Author name(s) |
| 2. | Year of publication |
| 3. | Study design |
| 4. | Country of publication |
| 5. | Age range of the study population |
| 6. | Sample size |
| 7. | Reported causes of mechanical small bowel obstruction (up to five) |

### Risk of bias assessment

Critical appraisal to assess the quality of the included studies will be carried out by two independent reviewers. We will use the Joanna Briggs Institute (JBI) tools (16) for each study design (case series, case-control, cohort) to be included in the review. We chose the JBI tools because they provide tools for a variety of study designs and are a widely used set of tools in systematic reviews. The JBI score is used to rate the quality of each included study as good, fair or poor. The quality rating would not be used to exclude a study from the review.

### Data synthesis

We anticipate that the eligible studies will be quite heterogeneous in terms of their design, outcomes of interest, populations and comorbidities. Therefore, results may be synthesised descriptively without meta-analysis. The characteristics of the included studies will be presented in a detailed table. In addition, important findings will be highlighted using charts and graphs. The review synthesis will be based on the major categories of small bowel obstruction aetiologies identified. Where possible, we will conduct a sub-analysis using conceptual frameworks based on age, WHO regions and continents.

## DISCUSSION

The proposed systematic review aims to examine the existing literature on the causes of mechanical bowel obstruction in LMICs from 2002 to 2022, with a particular interest in the common aetiology to guide surgical practice.

Identifying the evidence for common causes of mechanical bowel obstruction in the context of fragile health systems in LMICs over the last two decades is critical to halting the unacceptably high mortality associated with this surgical condition. This review has the potential to provide surgical trainees with scientific evidence to raise a high index of suspicion for prompt diagnosis in patients presenting with acute abdomen to the emergency department. Making the correct diagnosis would enable surgeons to offer appropriate interventions to patients in need, regardless of the limited diagnostic modalities in LMICs.

The review would identify gaps that could potentially be explored through further research by scientists to better understand the aetiology of mechanical bowel obstruction in LMICs. In addition, the evidence to be synthesised may also assist academics in communicating new knowledge, as well as policymakers in formulating appropriate policies that could improve surgical needs and outcomes.

The review will have some limitations. Because it is limited to LMICs, a real-time comparison with the more recent common aetiology of mechanical bowel in HICs would be difficult. However, the robust health systems in HICs, including advanced diagnostic modalities, can prevent missed diagnoses and prompt treatment. Furthermore, the aetiological pattern of mechanical bowel obstruction in this setting is unlikely to have changed, given the recent incontrovertible evidence from empirical studies. Furthermore, due to a lack of funding, librarians could not be consulted for expert input, which may have resulted in some important empirical studies being overlooked. Despite these limitations, this review, when completed, has great potential to add to the body of knowledge, particularly in the surgical community, to improve outcomes. It will also shape future research in the gaps that remain to be identified.

## Data Availability

No datasets were generated or analysed during the current study. All relevant data from this study will be made available upon study completion.

